# Breath-Based Monitoring of High Cholesterol State and Statin Therapy

**DOI:** 10.1101/2025.09.19.25336177

**Authors:** Ashok Prahbu Masilamani, Palash Kaushik, Mojtaba Khomami Abadi, Fatemeh Yazdanpanah, Jayden Hooper, Hélène Yockell-Lelièvre, Kim Sergerie, Anick Dubois, Frédéric Lesage, Jean-Claude Tardif

## Abstract

Monitoring the effectiveness of statin therapy in patients with dyslipidemia is essential for ensuring optimal treatment outcomes. The current standard involves lipid profiling via blood tests to detect abnormalities in blood lipids. This study evaluated the feasibility of a non-invasive, breath-based approach to statin therapy monitoring using Noze’s electronic nose (eNose) platform. A total of 35 participants were enrolled, 25 with elevated low-density lipoprotein cholesterol (LDL-C) levels and 10 healthy controls. The high LDL-C group provided breath specimens both before starting statin therapy and after 6 to 8 weeks of treatment. These breath specimens were digitized using Noze’s eNose platform and analyzed using machine learning (ML) algorithms. Results showed a 91% sensitivity and 87% specificity in identifying high blood cholesterol cases, demonstrating the potential of Noze’s eNose platform for non-invasive monitoring of statin therapy through exhaled breath.

## 1. Introduction

Cholesterol plays a fundamental role in vertebrate metabolism, serving as a key precursor for essential biological compounds such as bile acids, corticosteroids, sex hormones, and vitamin D-derived hormones ^1^. However, despite its physiological importance, an elevated cholesterol level—known as hypercholesterolemia—is a major risk factor for numerous cardiovascular diseases (CVDs), including coronary atherosclerosis, cerebrovascular disease, and peripheral artery disease ^2–4^. According to the World Health Organization (WHO), CVDs accounted for 17.9 million deaths globally in 2019 ^4^. Evidence indicates that cardiovascular risk can be mitigated by managing modifiable factors such as blood cholesterol, blood pressure, and tobacco smoking ^5^. Among these, cholesterol management is commonly addressed through the use of statins, which inhibit the enzyme 3-hydroxy-3-methylglutaryl-coenzyme A (HMG-CoA) reductase, a key regulator in the mevalonate pathway of cholesterol biosynthesis (Figure 1). Therefore, statins reduce the level of blood cholesterol ^6,7^.

**Figure 1.**
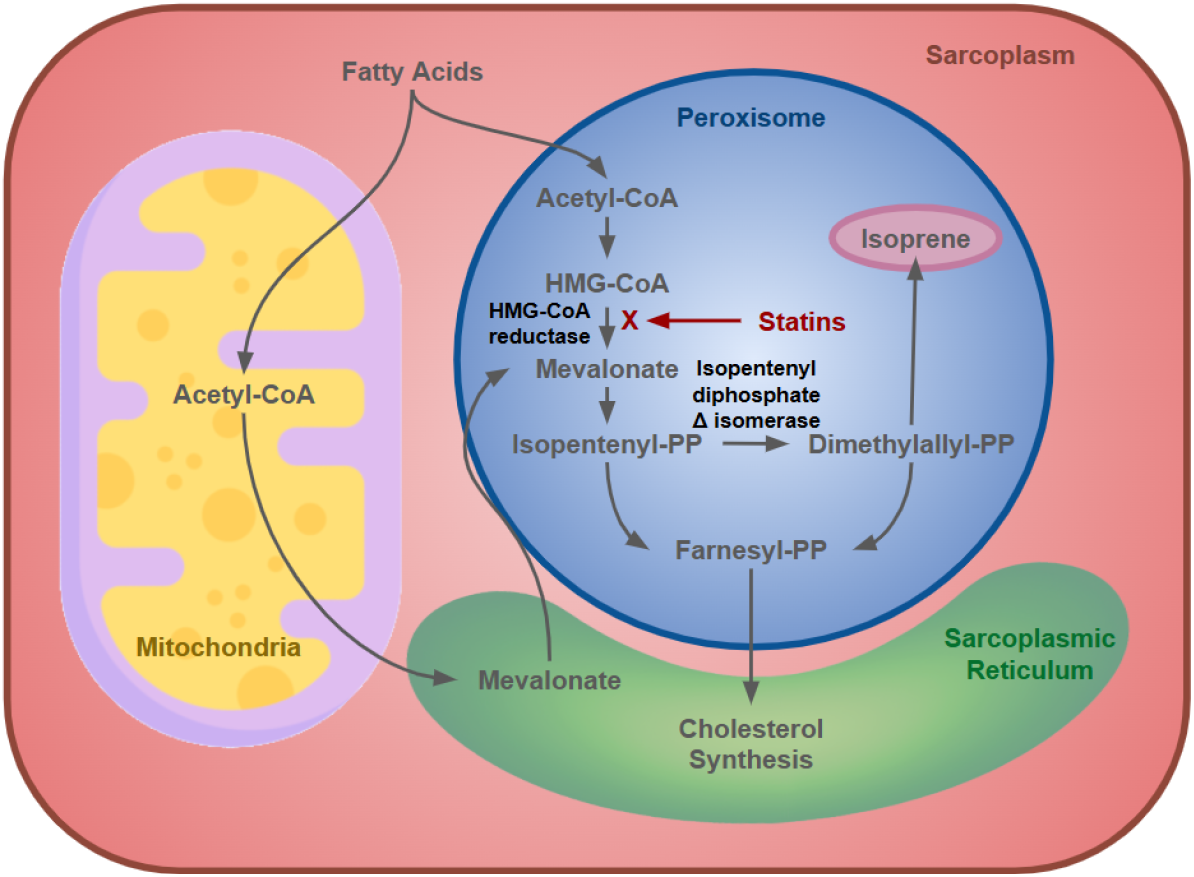
Depiction of the mevalonate pathway active in skeletal muscle tissue.

Effective cholesterol management with statin therapy involves individualizing both the statin type and dosage. A study conducted on an Asian patient population demonstrated that these factors significantly influence LDL-C reduction in targeted populations ^8^. Tailoring statin therapy to each patient can improve treatment outcomes, but it also necessitates cholesterol monitoring that is comfortable and convenient for patients. Recent advancements in breath analysis, particularly those based on electronic nose (eNose) technology, offer a non-invasive alternative for monitoring physiological changes ^5^. Breath analysis has already shown promise in clinical research for detecting respiratory infections ^9^, cancer ^10^, and other sources of metabolic disorders ^11,12^. eNose devices work by capturing the unique mixture of volatile organic compounds (VOCs) present in exhaled breath, generating a breathomic fingerprint (“breathprint”) that reflects the body’s metabolic health status. This emerging field of breathomics has gained considerable interest as a detection/ monitoring tool due to its non-invasive nature, ease of use, and capacity to deliver rapid insights into a patient’s health.

The application of an eNose device for detecting dyslipidemia via exhaled breath has traditionally been based on the detection of VOCs presumed to originate from cholesterol biosynthesis, particularly through the mevalonate pathway. This pathway, depicted in Figure 1, plays a central role in sterol production by converting acetyl-CoA, derived from glucose, fatty acid, and amino acid metabolism, into mevalonate via the action of HMG-CoA reductase ^13,14^. Mevalonate is then phosphorylated and processed into activated isoprenoid units, which are further transformed into squalene and subsequently lanosterol. Lanosterol undergoes multiple enzymatic steps to produce cholesterol. It has long been proposed that during this biosynthetic sequence, intermediates such as isopentenyl pyrophosphate (IPP) and dimethylallyl pyrophosphate (DMAPP) may degrade non-enzymatically into isoprene ^13,14^. By inhibiting HMG-CoA reductase, statins disrupt the mevalonate pathway responsible for cholesterol synthesis. This inhibition reduces the production of isoprenoid intermediates, thereby lowering isoprene formation. As a result, individuals with elevated blood cholesterol levels are expected to exhibit a significant decrease in breath isoprene levels following statin therapy.

However, recent literature has raised questions about the extent to which hepatic cholesterol biosynthesis contributes to exhaled isoprene levels, suggesting alternative or additional sources such as skeletal muscle peroxisomes may play a more prominent role ^15–17^. This suggests that VOCs other than isoprene, arising from lipid peroxidation, side-chain modifications, or downstream branches of sterol metabolism, possibly contribute to the breath profile associated with dyslipidemia. Oxidative transformations of sterol intermediates, cleavage of aliphatic side chains, or activity within parallel metabolic pathways such as bile acid synthesis or steroid hormone production may generate VOCs such as aldehydes, ketones, and short-chain hydrocarbons. These VOCs can diffuse into the bloodstream and reach the lungs, where they cross the alveolar membrane and are excreted into exhaled breath via pulmonary gas exchange.

The primary goal of this study was to evaluate the feasibility of using an eNose device prototype (Noze Inc., Montreal, Canada) as a non-invasive tool to monitor statin therapy in individuals with elevated blood cholesterol. Patients with high LDL-C levels from the Montreal Heart Institute were recruited for the study. The eNose device captured a digital “breathprint” (the digitized VOC profile of an individual’s breath) from each participant’s exhaled breath. By comparing breath profiles of participants with high blood cholesterol collected before and during statin treatment, the aim of the study was to establish that breathprints between the two conditions are distinctly identifiable.

The secondary objective was to analyze a subset of breath specimens from both groups using Gas Chromatography-Mass Spectrometry (GC-MS) to identify cholesterol-associated VOC biomarkers that show the greatest variation between untreated and treated individuals. This validation step aimed to provide biological plausibility for the breath pattern clustering detected by the eNose.

## 2. Materials and Methods

### 2.1 Study Design

This study employed a prospective, two-group design to evaluate the feasibility of using an eNose device for monitoring statin therapy. A group of participants with high LDL-C provided breath specimens before and after 6-8 weeks of statin treatment, allowing for a within-subject comparison. A separate control group with normal LDL-C levels provided a single breath specimen for comparison. The eNose data, analyzed using machine learning, aimed to distinguish between these groups and time points. Additionally, a subset of specimens underwent GC-MS analysis to identify potential VOC biomarkers associated with cholesterol levels and statin response, adding a validation component to the eNose study findings.

### 2.2 Study Population

The study enrolled 35 participants, divided into two cohorts. All participants underwent a complete lipid panel twice: once before the initiation of statin therapy, and a second time—only for the patient cohort—6 to 8 weeks after beginning statin treatment. Blood specimens were collected on the same day as breath specimens to ensure consistency. The patient cohort included 25 individuals with elevated LDL-C levels (>3.0 mmol/L), all of whom received oral statin therapy for 6 to 8 weeks. Each participant provided two sets of breath specimens: one prior to treatment and one following the treatment period. Each set consisted of 5 breath specimens delivered to Tedlar bags. The control cohort consisted of 10 participants with normal LDL-C levels (less than 3.0 mmol/L), who provided one set of breath specimens and one blood specimen.

### 2.3 Device Description

In order to identify a clear relationship between the metabolic processes and breath VOC profile, exclusion criteria were designed for patient selection. The exclusion criteria were tuned to limit the effect of metabolic activities associated with medical conditions that are unrelated to cholesterol biosynthesis. The exclusion criteria were: 1) use of statins at any point during the previous 6 months; 2) history of statin intolerance; 3) known or new-onset diabetes mellitus; 4) moderate to severe kidney disease with an estimated glomerular filtration rate of less than 45 ml/min/1.73 m2; 5) liver diseases such as cirrhosis, chronic active hepatitis, jaundice, hepatic insufficiency, transaminase levels greater than 3 times the upper limit of normal and/or total bilirubin levels greater than 2 times the upper limit of normal; 6) history of alcohol and/or drug abuse. To further limit confounding factors associated with metabolic activities, participants were instructed to fast for a minimum of 8 hours before breath specimen collection.

#### 2.3.1 eNose Device

Breath specimens from all 35 participants across both cohorts were sampled, using the Noze eNose device, a modular, cloud-connected system driven by Noze’s proprietary aroma chip technology. As shown in Figure 2a, the device comprises two primary components: the aroma chip module and the IOT module. At the heart of the aroma chip module is Noze’s proprietary sensor, equipped with a miniature integrated pump for specimen collection. The aroma chip, shown in Figure 2b, features an array of 32 chemiresistive receptors configured as thin films. Each receptor exhibits cross-sensitivity to various families of VOCs, enabling broad-spectrum chemical detection. Once powered on and activated, the eNose device collects a breath specimen from the Tedlar bag via the inlet tube illustrated in Figure 2d. The aroma chip module immediately processes the specimen, converting it into a digital breath fingerprint, referred to as breathprint. This data is then securely transmitted and stored in Noze’s cloud-based aroma breathprint database through the IoT module.

**Figure 2.**
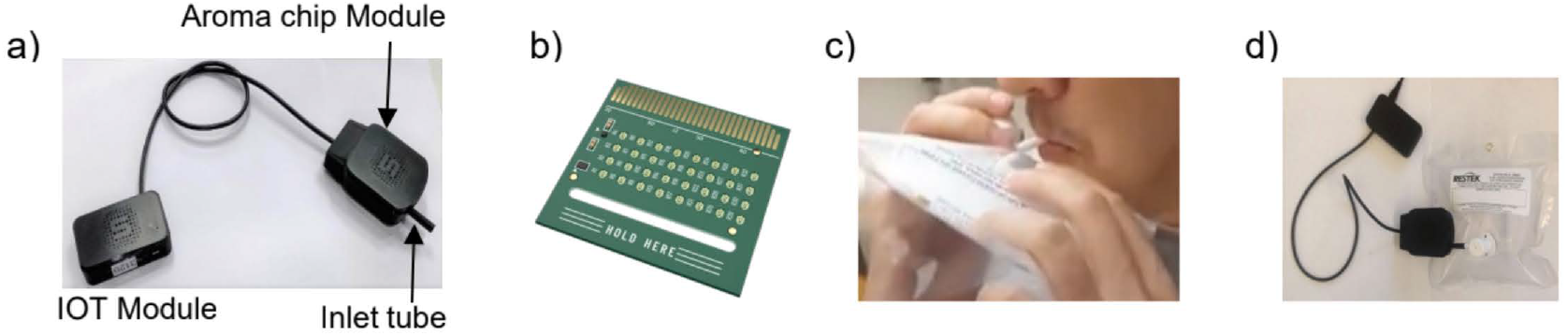
(a) Noze eNose device, (b) The aroma sensor chip, (c) Breath specimen being collected from a participant breathing into a Tedlar bag, (d) Breath specimen being introduced to the aroma chip module.

#### 2.3.2 Breath Specimen Digitization

Breath specimens were processed by the eNose device through a three-step procedure. In the first step (baselining), the device sampled ambient air from the surrounding environment for 60 seconds to establish a reference. Immediately following this, in the second step, the outlet of the Tedlar bag containing the breath specimen was connected to the device’s inlet tube (as shown in Figure 2d), allowing the aroma chip to analyze the specimen for 120 seconds. In the third step, the recovery phase, the Tedlar bag was disconnected, and the aroma chip was given 300 seconds to return to its environmental baseline.

When exposed to a volatile mixture such as a breath specimen, the impedance of the thin film receptors on the aroma chip sensor array is modified. The collective response of the 32 chemiresistive elements generates a unique digital breathprint in the form of a time series unique to the volatile mixture introduced. For each alveolar breath specimen collected in the Tedlar bag and sampled by the eNose device, a corresponding digital breathprint was generated. As illustrated in Figure 3, the plot displays the normalized impedance responses of all 32 sensor elements (s1 to s32) over time. Each participant’s breathprint can be interpreted as a 32-dimensional time series comprising three distinct phases: 1) the baseline, 2) the breath specimen introduction, and 3) the recovery.

**Figure 3.**
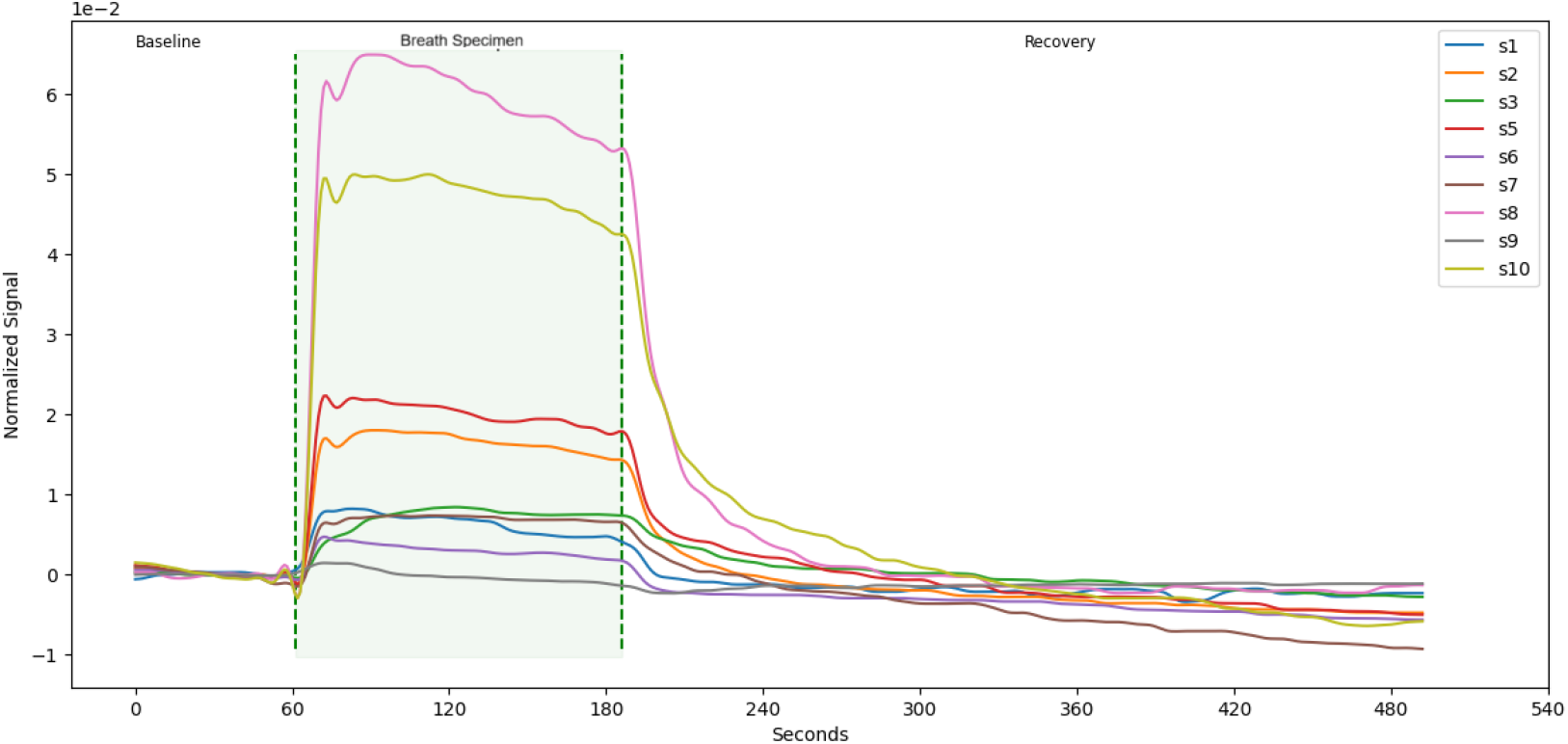
Time series generated by Noze’s aroma chip in response to exposure to exhaled alveolar breath. The signal plot is divided into three distinct phases: baseline (sampling ambient air), breath sampling (introduction of the breath specimen), and recovery (return to baseline conditions).

### 2.4 Breath Collection Procedures

#### 2.4.1 Breath Specimen Collection

Exhaled Breath was collected using 0.5L Tedlar breath bags (Restek), as shown in Figure 2c. Each participant provided five breath specimens. Before sampling, participants were instructed to relax, normalize their breathing, and perform three practice exhalation cycles to acclimate to the environment. During the actual collection, a specialized breathing procedure was used to exclude the dead space air; only the alveolar portion of the exhaled breath was captured in the bag. The specialized procedure comprised a series of three deep breathing cycles, followed by a fourth cycle wherein participants exhaled the last portion of their exhaled breath into a Tedlar bag.

#### 2.4.1 Breath Specimen Collection for GC-MS Analysis

Breath specimens from four participants in the patient cohort, collected prior to statin therapy, and from four others following statin administration were selected for analysis using a GC-MS system. One participant provided specimens at both time points. During each sampling session, two breath specimens were collected per participant. The specimens were transferred from Tedlar bags to Tenax-TA/Carbotrap dual-bed thermal desorption (TD) tubes (Markes), stored at temperatures between 0 °C and 4 °C, and shipped to a third-party lab partner for GC-MS analysis. All analyses were completed within three weeks of specimen collection.

### 2.5 Statistical and Data Analysis Methods

#### 2.5.1 Feature Extraction

The transient responses of the aroma chip, shown in Figure 4, reflect impedance changes in the sensor’s thin films during the three phases of the breath sampling process. These normalized time series capture the dynamic behavior of the sensor array when exposed to exhaled breath, enabling the extraction of multiple engineered features that characterize VOC interactions. Key features include: 1) Extremum of the transient response: represents the maximum deflection in impedance, corresponding to the greatest expansion of a thin film during the physical absorption of VOCs with high affinity to the receptor. 2) Area under the curve (AUC): captures both the extent of VOC absorption and the rate of physical changes in the sensor film during absorption and desorption. A larger AUC may indicate a rapid absorption, suggesting strong VOC affinity and quick film expansion, and/or a slow desorption, also implying high affinity, as the film takes longer to return to baseline. 3) Physical absorption rate: the impedance change during the first 10 seconds of specimen exposure is measured as a point-to-point slope, providing insight into the presence and concentration of VOCs based on the initial film expansion rate. 4) Normalization with respect to humidity: Given that the sensor thin films also have affinity for water vapor and that breath humidity levels can vary, a complementary set of features was engineered by normalizing the above metrics against the maximum change in relative humidity during the sampling phase (relative to baseline). This step ensures robustness of VOC signal interpretation despite variations in breath moisture content.

**Figure 4.**
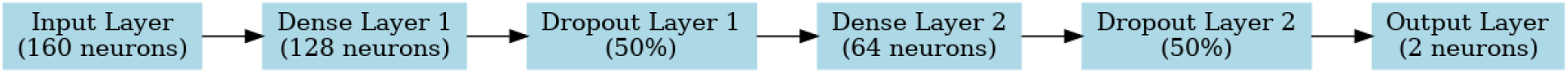
Multilayer perceptron neural network model that was trained on the dataset.

For each digitized breath specimen, engineered features are extracted from the transient response during the breath sampling phase, resulting in a 160-dimensional vector that serves as the specimen’s descriptor. Each descriptor is uniquely labelled based on the participant’s ID and categorized by cohort and subtype—control, pre-statin, or post-statin. Further details on data processing can be found in the supplementary file.

#### 2.5.2 AI Processing

A multilayer perceptron (MLP) neural network, illustrated in Figure 4, was trained using two hidden dense layers with 128 and 64 neurons, respectively. Each neuron employed the ReLU activation function, and both layers were followed by dropout layers with a dropout rate of 0.5 to enhance model generalization. The dataset was randomly split into training and validation sets, with 80% allocated for training. Model weights were optimized using the Adam optimizer. Given the inclusion of participant-specific information in the data, the MLP was designed to achieve two primary objectives: (i) preserve task-relevant information for distinguishing between pre-statin and post-statin breath specimens, and (ii) minimize the influence of participant-specific features irrelevant to the classification task. Additionally, the model was trained to learn a vector space representation that reflects the semantic structure of the data while reducing inter-participant variability.

The model architecture was intentionally kept shallow, with only two hidden layers, to prevent overfitting and ensure interpretability. ReLU was selected as the activation function to maintain the model as a piecewise linear approximator.

### 3. Results

### 3.1 Descriptive Statistics

During the study, twenty-five subjects were enrolled in the patient cohort, and ten subjects comprised the control cohort. The mean pre-statin LDL-C level for the patient cohort was 4.04 mmol/L, which decreased to 1.99 mmol/L following six to eight weeks of statin therapy. Table 1 provides a summary of the participants’ clinical characteristics.

**Table 1.** Demographics and clinical characteristics of the study population.

| Parameter | Healthy controls<br>mean ± SD | Patients<br>mean ± SD |
| --- | --- | --- |
| Number of participants | 10 | 25 |
| Sex | Female - 4<br>Male - 6 | Female - 11<br>Male - 14 |
| BMI mean (kg/m<sup>2</sup>) | 24.65 ± 2.80 | 27.76 ± 4.92 |
| Pre-statin LDL-C level (mmol/L) | 2.05 ± 0.76 | 4.04 ± 0.87 |
| Post-statin LDL-C level (mmol/L) | - | 1.99 ± 0.73 |

### 3.2 Statistical Analysis of Descriptor Distributions

The inter-participant and intra-participant variability of the digitized breath specimens was assessed by comparing the average distance between breath specimen descriptors across individuals. The ratio of intra-to inter-participant variance was calculated, yielding a mean of 3.0% and a standard deviation of 1.5%. This relatively low variance ratio indicates that breath specimens from the same individual are more similar to each other than to those from other participants. This outcome reflects the influence of individual-specific factors, such as dietary habits, physiological differences, and microbiome composition, on breath VOC profiles, in addition to cholesterol levels.

Each patient participated in two breath sampling sessions—one prior to and one following statin therapy. The ratio of intra-session variance (within the same participant) to inter-session variance was calculated, yielding a mean (μ) of 1.4% and a standard deviation (σ) of 0.8%. This low ratio suggests that breath specimens from different sessions of the same individual are distinguishable. These findings indicate that the digitized breath data captures both (i) task-specific information relevant to detecting changes in cholesterol levels due to statin therapy, and (ii) participant-specific characteristics.

### 3.3 AI Model Output

During the training process, specimens from the control group were assigned the same class label as those from the post-statin group. To address potential class imbalance caused by the overrepresentation of the normal-cholesterol category, a weighted binary cross-entropy loss function was employed for training the MLP. Early stopping was applied to prevent overfitting and ensure that the model remained generalizable to the validation dataset. This approach yielded a sensitivity of 91% and specificity of 84% for detecting the high-cholesterol class (pre-statin) when the control group was excluded. Upon inclusion of the control group data, performance improved, achieving 91% sensitivity and 87% specificity.

To visualize the data representation underlying the model’s performance, descriptors were extracted from the output of the second hidden dense layer, resulting in a 64-dimensional vector space referred to as the model’s representation space. To enable two-dimensional visualization of this high-dimensional space, the Uniform Manifold Approximation and Projection (UMAP) technique was applied which is a widely used unsupervised dimensionality reduction method. This approach illustrates the distribution of breath specimens as learned by the model for the classification task. In the resulting plots (Figure 5), each digitized breath specimen is represented as a dot.

**Figure 5.**
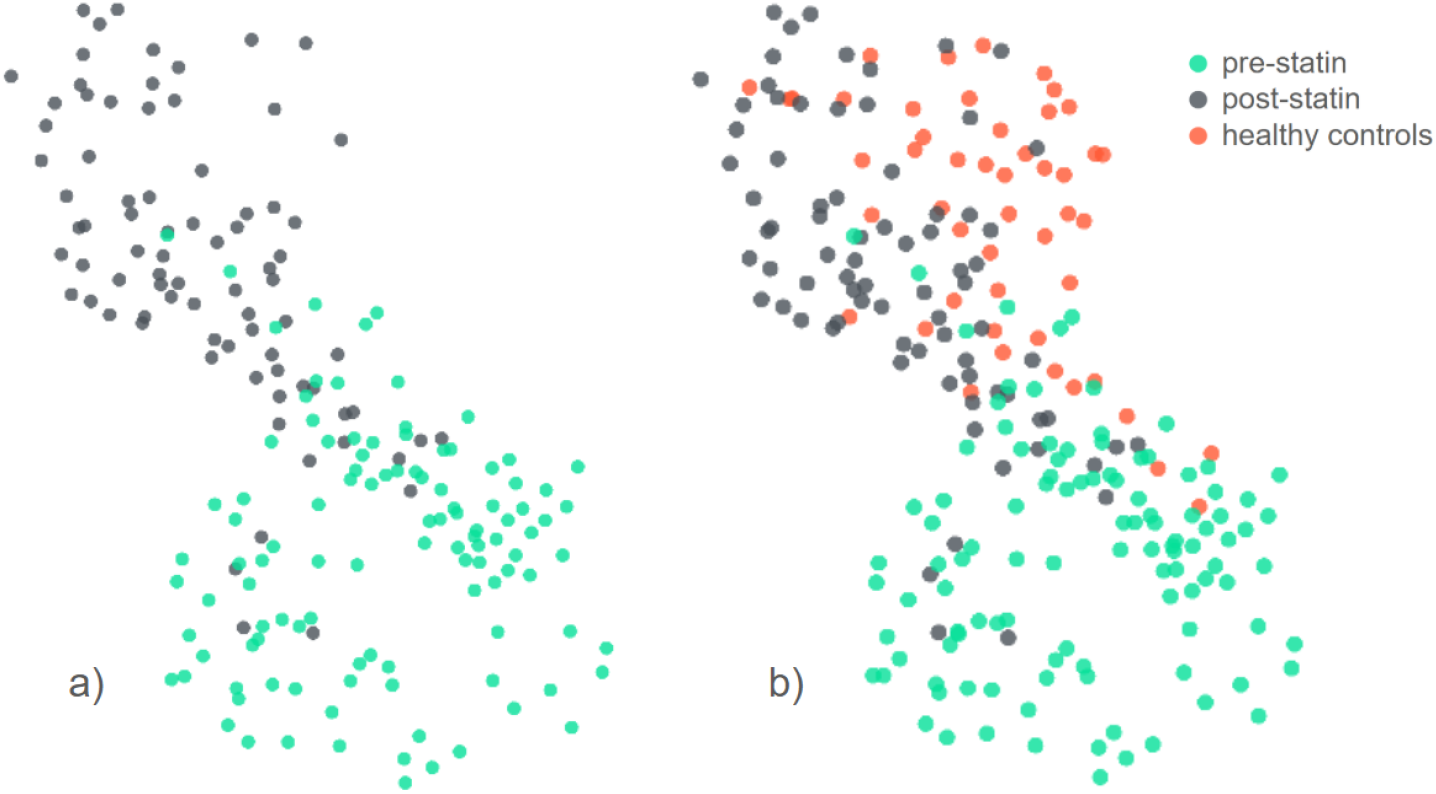
Two-dimensional plots of breath specimen data visualized using UMAP; (a) breath specimens from participants in the patient cohort, collected before and after statin therapy; (b) breath specimens from the control cohort plotted within the same representation space as the patient cohort.

Through the analysis conducted via GC-MS, it was possible to isolate the concentration of isoprene from the breaths of participants from the patient cohort before and after statin therapy. Figure 6 displays the measured breath isoprene levels and the corresponding LDL-C levels from the same patients at the time of sampling.

**Figure 6.**
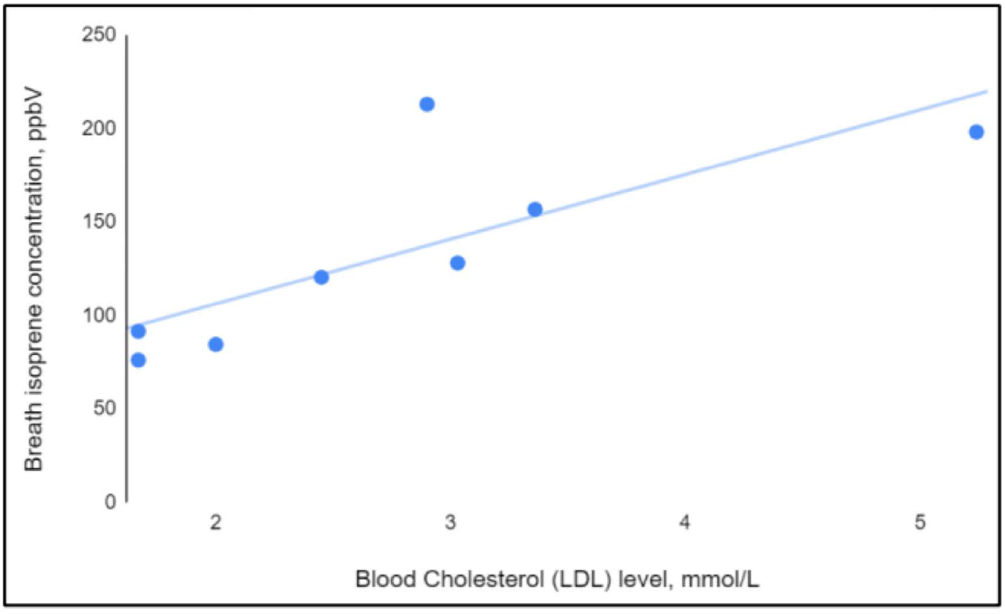
Blood LDL-C levels and corresponding breath isoprene levels for 8 participants (patient cohort) of the study.

## 4. Discussion

The VOC profile captured by the eNose device effectively discriminates individuals with elevated LDL-C levels prior to statin therapy from both post-therapy individuals and healthy controls, the latter two displaying overlapping breath signatures. Complementary analysis using GC-MS identified a statistically significant correlation between blood LDL-C concentrations and exhaled isoprene levels (Figure 6). Specifically, the Pearson correlation coefficient between blood cholesterol and breath isoprene was calculated at r = 0.785, with a corresponding p-value of 0.021, indicating a strong positive linear relationship that is unlikely to have occurred by chance. While recent studies have demonstrated that isoprene does not originate from hepatic cholesterol biosynthesis, as previously assumed, but rather from muscular lipid metabolism via the IDI2-dependent pathway ^16,17^, the observed correlation may reflect broader systemic effects of lipid-lowering interventions.

Statin therapy, by modifying lipid homeostasis, may influence skeletal muscle metabolism, oxidative stress, or mitochondrial activity, thereby altering isoprene output. Moreover, the overall breath signature captured by the eNose likely reflects a composite of multiple VOCs beyond isoprene alone. Statins can affect a wide range of metabolic pathways, including those involved in inflammation, bile acid synthesis, and oxidative processes, each of which may contribute to changes in the breath volatilome. A study by Henderson et al. ^18^ investigated the impact of statin use on exhaled breath VOCs during prolonged exercise. The research found that statin users exhibited different breath VOC profiles compared to non-statin users, particularly in the concentrations of short-chain fatty acids (SCFAs) like acetic acid, butanoic acid, and propionic acid ^18^. These differences suggest that statin metabolism may influence VOC emissions, possibly through effects on gut microbiota activity and associated metabolic processes. Furthermore, the study observed that isoprene levels in breath decreased with exercise and that this trend was more pronounced in statin users. Isoprene is a byproduct of the mevalonate pathway, which is inhibited by statins, leading to reduced cholesterol synthesis. Therefore, the decreased isoprene levels in statin users’ breath may reflect the pharmacological action of statins on this metabolic pathway. These multi-VOC shifts may underlie the observed clustering of post-treatment patients with healthy controls, and support the potential of breath analysis as a non-invasive integrative readout of systemic metabolic status.

## 5. Discussion

This study demonstrates that breath analysis using Noze’s eNose device can effectively distinguish between individuals in pre- and post-statin therapy states, and by extension, between elevated and reduced blood LDL-C levels. Complementary GC-MS analysis identified isoprene as strongly associated with these metabolic changes. Isoprene levels exhibited significant variation between pre- and post-statin samples and showed a strong correlation with blood LDL-C levels, supporting its role as a marker of endogenous cholesterol biosynthesis. Further analysis of the GC-MS data revealed a consistent relationship between LDL-C concentration and isoprene levels in breath. Also, collective analysis showed that breath specimens from individuals who underwent statin therapy were similar to those from healthy controls, when compared to pre-statin specimens. This suggests that differences between the breath VOC profiles of pre and post statin samples are not likely due to statin metabolism. This supports the potential of isoprene as a non-invasive biomarker for monitoring systemic lipid metabolism, especially under conditions where muscular cholesterol turnover or peroxisomal activity is altered. Additionally, the broader breath VOC breathprint captured by the eNose suggests that statin therapy and cholesterol regulation influence other volatiles beyond isoprene, which contribute to the discrimination of metabolic states. Collectively, the strong correlation between exhaled VOC profiles and blood lipid biomarkers underscores the potential of Noze’s eNose device as a non-invasive tool for monitoring cholesterol dynamics in both clinical and decentralized healthcare contexts.

## Data availability statement

The data that support the findings of this study are available from the authors upon reasonable request.

## Funding

This clinical pilot study was supported by the Health Collaboration Acceleration Fund from the Government of Quebec, as part of the project titled “*Paradigm Shift in the Conduct of Clinical Trials*”.

## Conflict of interest

Ashok Prabhu Masilamani, Palash Kaushik, Mojtaba Khomami Abadi, Fatemeh Yazdanpanah, Jayden Hooper, Helene Yockell-Lelievre, Kim Sergerie are employees of Noze Inc.

## Ethical statement

The clinical pilot study was approved by the Institutional Review Board of the Montreal Heart Institute (#2023-3132) and conducted in accordance with the principles embodied in the Declaration of Helsinki. All participants provided written informed consent prior to enrolment.

