## Supplemental Information for "Breath-Based Monitoring of High Cholesterol State and Statin Therapy"

### Supplementary Information

#### Data Processing

***Data yield and sample filtering**:* Data Yield - The yield of the specimens was majorly affected by three factors: internet connectivity, device failure and protocol adherence. These three factors impacted 10% of the data yield and hence total yield of the data collected by Noze eNose device was 90%; specifically, 5% reduction in yield occurred due to network connectivity issues and device and protocol adherence contributed to the other 5%.

***Digitized Breath Data Schema**:* Each digitized breath recording captures the transient responses of the Noze Aroma chip's chemical receptors, along with the transient responses of a temperature and relative humidity sensor, corresponding to one complete digitized breath. The transient responses are sampled at a frequency of 1 Hz and organized in a tabular format, with rows representing sequential timestamps and columns representing measurements from the transient responses. Each breath specimen recording consists of a 1-minute ambient baseline recording, a 2-minute breath bag sampling period, and a 5-minute device recovery period.

***Data Recording Validation**:* Prior to processing the information content of the digitized breath data, each digitized breath specimen was validated to ensure complete recordings and continuous transient response capture at 1 Hz, without data gaps due to potential network connectivity issues.

***Time Series Processing**:* The sensor thin films in the aroma chip are subject to drift over time due to factors such as temperature fluctuations, humidity variations, and aging. Drawing inspiration from computational neuroscience, to assess the information content of induced changes in the aroma chip's transient responses, the specimen exposure-induced transient responses are normalized against the sensor thin film's ambient state measures (baseline). The average measurements of the transient response during the baseline interval are calculated as the ambient baseline reference, denoted as R0. Normalized transient responses are calculated by subtracting R0 from the measured responses, and then dividing the result by R0.
